# Individual Preparedness for Distant Wildfires and the Delta Variant in the United States: A Survey of 2,250 US Residents

**DOI:** 10.1101/2021.07.24.21260660

**Authors:** Devabhaktuni Srikrishna

## Abstract

**Background:** COVID-19 virus travels in the air and collects indoors through tiny particles from exhaled breath, and remains a growing concern globally especially since case studies of vaccine breakthrough infections are being reported. Last year’s wildfires resulted in the worst air quality on record in the Western US due to toxic wildfire smoke (PM 2.5 pollution) traveling from distant wildfires and this year can potentially be even worse due to extremely dry conditions. Aerosol precautions such as high-filtration (Hi-Fi) masks and HEPA air purifiers are useful to effectively reduce inhalation of most of these toxic aerosols. Whereas the lack of fit or filtration in a mask or use of an air purifier of insufficient size (capacity) for the room can inadvertently render these precautions ineffective. Here we investigate the public’s concerns about wildfires and the COVID-19 variants (e.g. delta), their use of aerosol precautions, and whether these are being done in an effective manner.

**Methods:** We conducted a national survey of 2,250 US residents in order to understand public concerns about airborne threats and their usage of airborne (aerosol) precautions.

**Results:** We find over 66% of US residents surveyed are worried about inhaling COVID-19 and its variants, and 52% are worried about toxic wildfire particles in the air. In the mountain and pacific regions the latter rises to 73%. Only a quarter are using masks with higher filtration and high level of fit (or Hi-Fi masks e.g. N95 or similar such as elastomeric N95 or KF94). Two-thirds are still using loose-fitting cloth or surgical masks. Just over 40% of respondents report using air purifiers at home, and of this group only 40% use it in their bedroom where they sleep. Of those using air purifiers, the majority said they chose the size of their air purifier based on “most popular” models, “recommendations,” or “reviews.” However, of those using air purifiers only 42% reported doing a calculation (or using a calculator) to estimate the right size of air purifier needed for the room they are using it in. Notably, a much higher percentage of people (than average) reported use of Hi-Fi masks and home air purifiers in certain occupations such as doctors, healthcare, first responders, public safety, engineering, military, and construction.

**Conclusion:** National survey data suggests most US residents are worried about wildfire smoke and Covid variants (e.g. delta variant) but a majority are not prepared for it. Preparation with aerosol precautions will also be useful for future pandemics and national biodefense.

## Background

COVID-19 mutations such as the delta variant are becoming the greatest threat to national and global efforts to control the pandemic, as case studies of vaccine breakthrough infections are being reported. COVID-19 (virus) travels in the air and collects indoors [1]. The WHO, Los Angeles, San Francisco, and Israel have all recommended masks even for fully vaccinated people as case studies of vaccine breakthroughs [2] are being reported among fully vaccinated people and exhaled breath can contain COVID-19 and its variants (e.g. delta B.1.617.2) [3]. Preliminary studies suggest the people infected with the delta variant have a thousand times more virus in their respiratory tracts than with the original strain [4]. Even fully vaccinated people had a significant risk of infecting their unvaccinated household members in a study from Israel [38].

Evidence in support of aerosol transmission [5] continues to increase. Although contact with shared surfaces cannot be completely ruled out, it is a more likely explanation than droplets for the spread [6] of the virus in a large study in which patients in shared hospital rooms were infected in spite of being more than six feet apart and also separated by curtains. Whereas in a UK hospital, upgrading to FFP3 respirators (aerosol filtering) from loose-fitting surgical masks almost eliminated infection risk [7].

At the same time, the Western US is experiencing record breaking temperatures and extremely dry conditions predicting an earlier and more severe wildfire season than last year which resulted in the worst air quality on record in this region. Wildfire-specific pollution may be much more toxic [8] than other sources, particularly for young children [9] and people with pre-existing medical conditions [10], e.g. asthma, COPD and heart disease. Particulate pollution levels (PM aerosols) can be tracked in real-time locally [11] [12] or individually using home meters [13].

## Materials and Methods

### Survey Description

Surveys were administered between June 30 through July 7, 2021 via web platform with demographic information including income and occupation. Surveys queried respondents’ level of concern about pollution from distant wildfires and COVID-19 variants, their choice of masks (e.g. cloth versus N95) and air purifiers, their planned usage of masks and purifiers in the next six months in different environments (e.g. bedroom, home, school, office), and whether they actually conducted a calculation to choose the size of their air purifier for the room they use it in. More than 93% of those who started taking our survey completed it. The respondents themselves are not paid. SurveyMonkey reaches out to their audience and respondents get to choose a charity of choice to receive a donation for doing the survey. The survey aims to be reasonably balanced across regions, gender, ages, and incomes.

- Sample Size: 2250 total completed responses from US residents
- Regions: 28.5% from pacific/mountain, 35.5% East Coast, 33% Central
- Gender: 57% female / 43% male
- Ages: 27.5% 18-29 years, 24.5% 30-44 years, 29% 45-60 years, 19% > 60 years
- Annual Household Income: 40% < $50k, 30% $50k-$100k, 20% > $100k, remaining prefer not to answer

### SurveyMonkey platform

The web-based survey was hosted on the SurveyMonkey.com platform and was deployed using SurveyMonkey’s audience panel [14]. SurveyMonkey is a cloud platform on which respondents complete surveys. SurveyMonkey has a pool of millions of respondents who have volunteered to take surveys. SurveyMonkey recruits respondents from their pool to take a particular survey [15].

Once notified, volunteers have the choice to opt-in to take any of their surveys they choose (and become respondents). There is no obligation for the respondent to take the survey because respondents volunteered for SurveyMonkey. The incentive for the respondent is if they complete a survey, then SurveyMonkey makes a small donation ($0.50) to a charity designated by that respondent. Donations are to the charity chosen by the respondent (not to respondent directly) and are made for completing the survey all at once not on a per question basis.

Respondents have no arrangement with the author, nor is their identity known to the author. SurveyMonkey is paid directly by the author. PatientKnowhow, Inc. sponsored the survey and has no relation to any of the manufacturers discussed.

### Level of concern about inhalation of Coronavirus and toxic wildfire particles

To understand how concerned people are about aerosol inhalation risks, survey respondents were asked questions on a five point scale about how worried they are about inhaling Coronavirus and its variants during the next six months (e.g. Delta variant), from “a great deal,” “a lot,” “a moderate amount,” “a little,” or “none at all.” Similarly they were asked how worried they are about inhaling particulate pollution from wildfires (e.g. PM 2.5) during wildfire season (July-November).

### Frequency and type of mask used

The type of mask primarily used by respondents can be broadly classified into two types: Hi-Fi (those with high-filtration and good fit to form a face seal) and everything else. To understand the utilization of Hi-Fi masks that filter aerosols, survey respondents were asked questions about how often they expect to use a mask to protect themselves from wildfire particulates or COVID-19 (and its variants) in the air in the next 6 months on a five point scale, “never,” “rarely,” “monthly,” “weekly,” and ‘daily.” They were also asked what is the primary mask they use to protect themselves from Coronavirus (Covid-19) with multiple choices including cloth masks, surgical masks, N95 masks, and several other choices detailed in the results section.

### Frequency, location, and size calculation of air purifier used

To understand the usage of air purifiers and HEPA air cleaners, Survey respondents were asked questions about how often they expect to use an air purifier or HEPA air cleaner in the next 6 months on a five point scale, “never,” “rarely,” “monthly,” “weekly,” and “daily.” They were also asked the location where they currently use an air purifier or HEPA air cleaner or plan to during wildfire season with multiple choices including at home, in the bedroom where they sleep, in their living area during the day, at work, and at school. Finally they were asked how they decided the size of air purifier or HEPA air cleaner need in the room they use it in with five choices including

- “Not applicable”
- “I did not do a calculation, but I chose a popular model”
- “I did a calculation myself using clean air delivery rate (CADR) and dimensions of the room (length x width x height)”
- “I used a calculator (e.g. online) designed for this purpose by inputting the dimensions of the room (length x width x height)”
- “I did not do a calculation, but took the advice of a friend or colleague”
- “I did not do a calculation, but took the advice of a review or article”

### Profession

To correlate the above usage choices of masks and air purifiers by occupation, survey respondents were asked about their profession with multiple choices spanning a variety of industries as detailed in the results section.

## Results

Concerns about aerosols and use of precautions were evaluated using N=2,250 survey responses from across the country whose approximate locations are shown in Figure 1. These represent a broad cross section of the United States from Western, Mountain, Central, and Eastern time zones with higher density of responses reflective of population centers.

**Figure 1:**
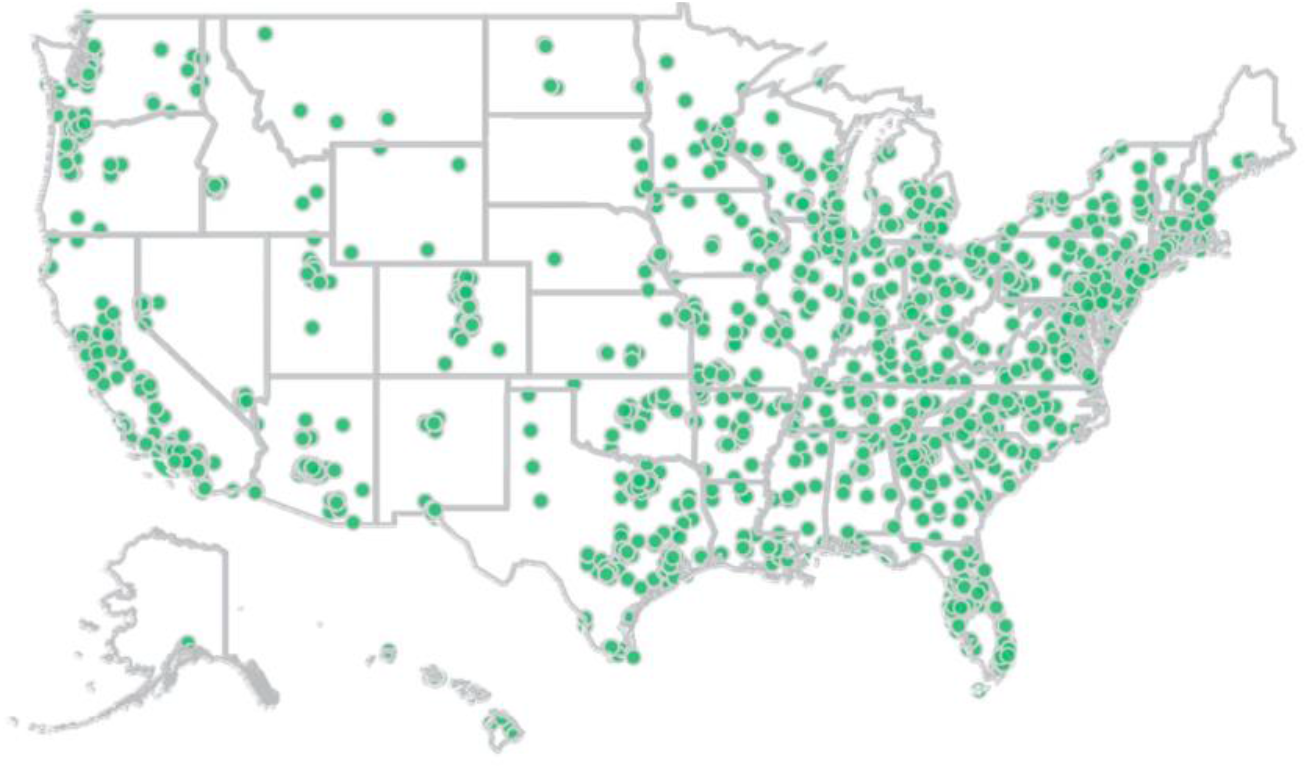
Approximate locations of N=2,250 survey respondents across the United States.

### The majority of US residents are worried about Coronavirus and toxic wildfire particles

Respondents’ concern for inhaling aerosols from COVID-19 (and its variants such as delta) and wildfire pollution was measured on a five point scale as shown in Figure 2. Approximately 66% of respondents across the country reported being moderately or more worried about inhalation of COVID-19 and about 18% worry about it “a great deal.”

**Figure 2:**
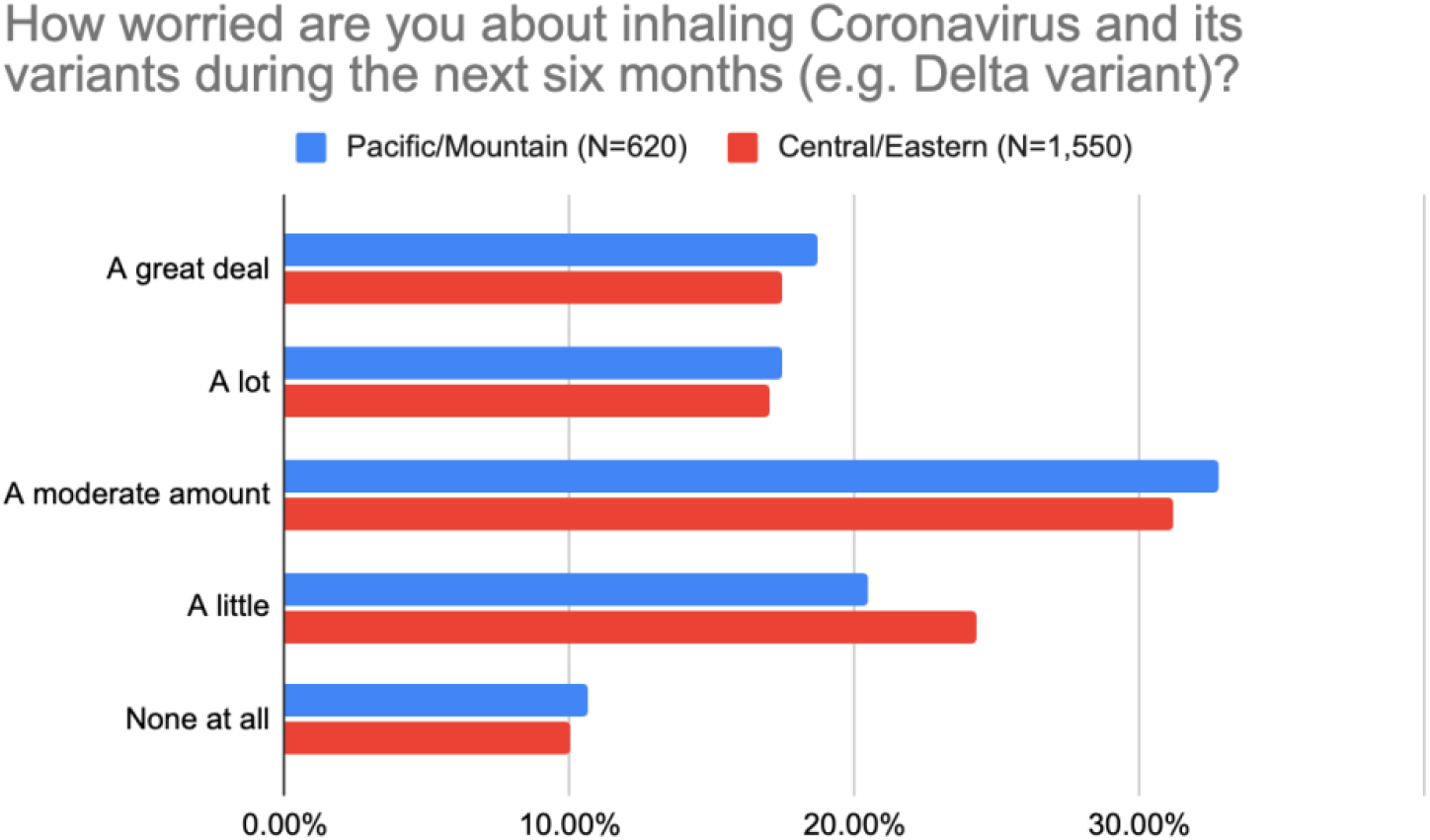
Concerns of US residents about Coronavirus and its variants (e.g. Delta variant)

However there was significant variation between parts of the country with respect to worry about wildfire pollution consistent with the regional risk in the Western United States (Pacific/Mountain) as shown in Figure 3. Only 42% of respondents expressed moderate or more worry about wildfire smoke in the Central/Eastern parts whereas 73% did so in the Pacific/Mountain areas with 19% worried about it a “great deal.” Nationally 52% of respondents expressed moderate or more worry about particulate pollution from wildfires.

**Figure 3:**
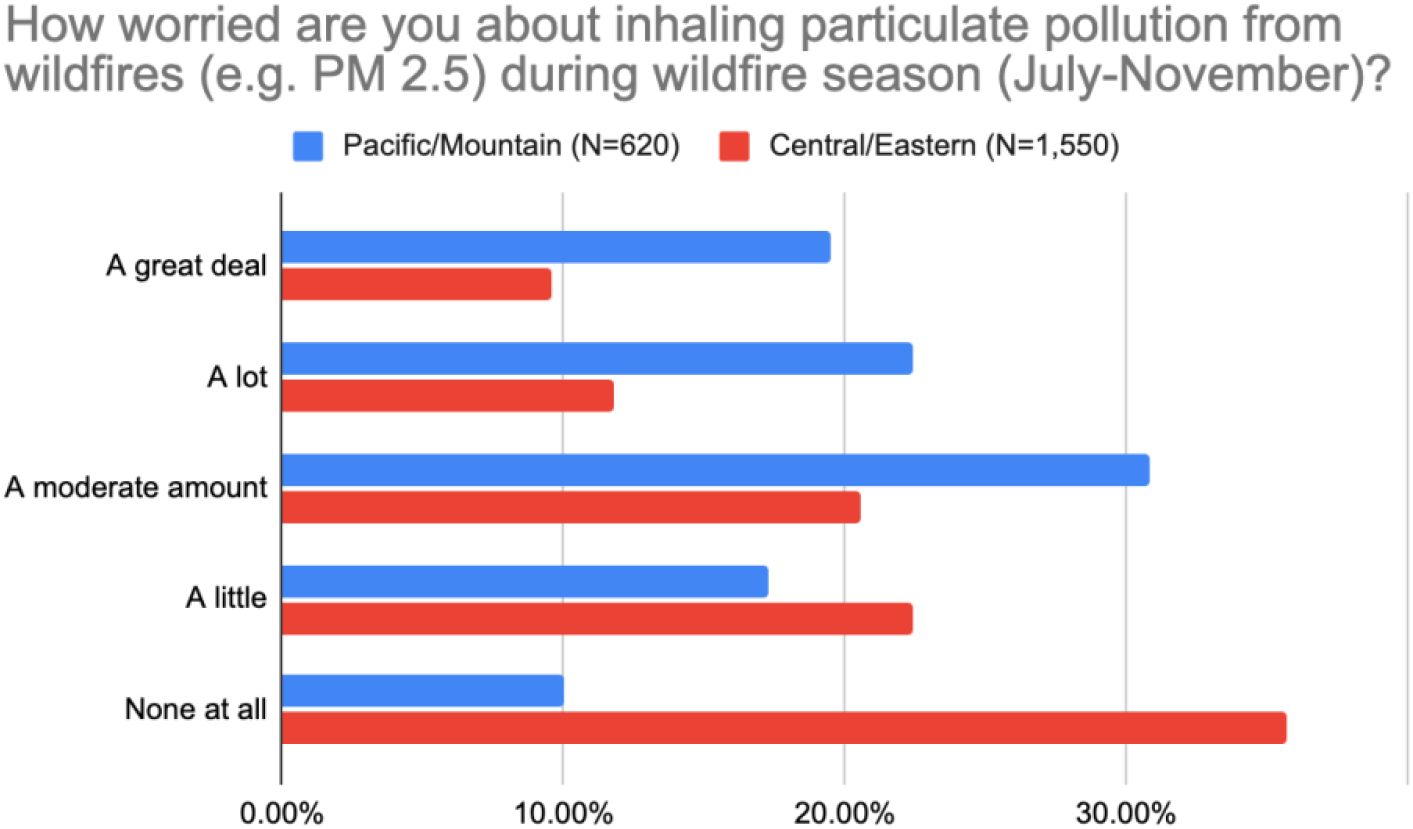
Concerns of US residents about particulate pollution from wildfires.

### Only a quarter of US residents use Hi-Fi masks but it varies by occupation

Respondents were asked which masks they primarily use and the percentage using Hi-Fi masks varies by profession. Although there are significant differences in quality, the Hi-Fi masks listed as multiple choices in this survey include disposable N95 masks or similar such as elastomeric N95, KF94, surgical masks with fitters, KN95 masks, and so on. In contrast, loose fitting masks and those with low filtration include surgical masks without fitters, cloth masks, gaiters, or no mask at all.

Overall, a quarter or 27% of respondents reported using Hi-Fi masks as their primary mask, however this percentage varied between 15% and 50% based on the occupation of the respondent. As shown in Figure 4, the occupations with highest percentage of Hi-Fi masks are perhaps not surprisingly first responders, doctors, nurses, and firefighters for whom these are likely to be used at work but curiously also among those in the restaurant industry, military, construction and engineering.

**Figure 4:**
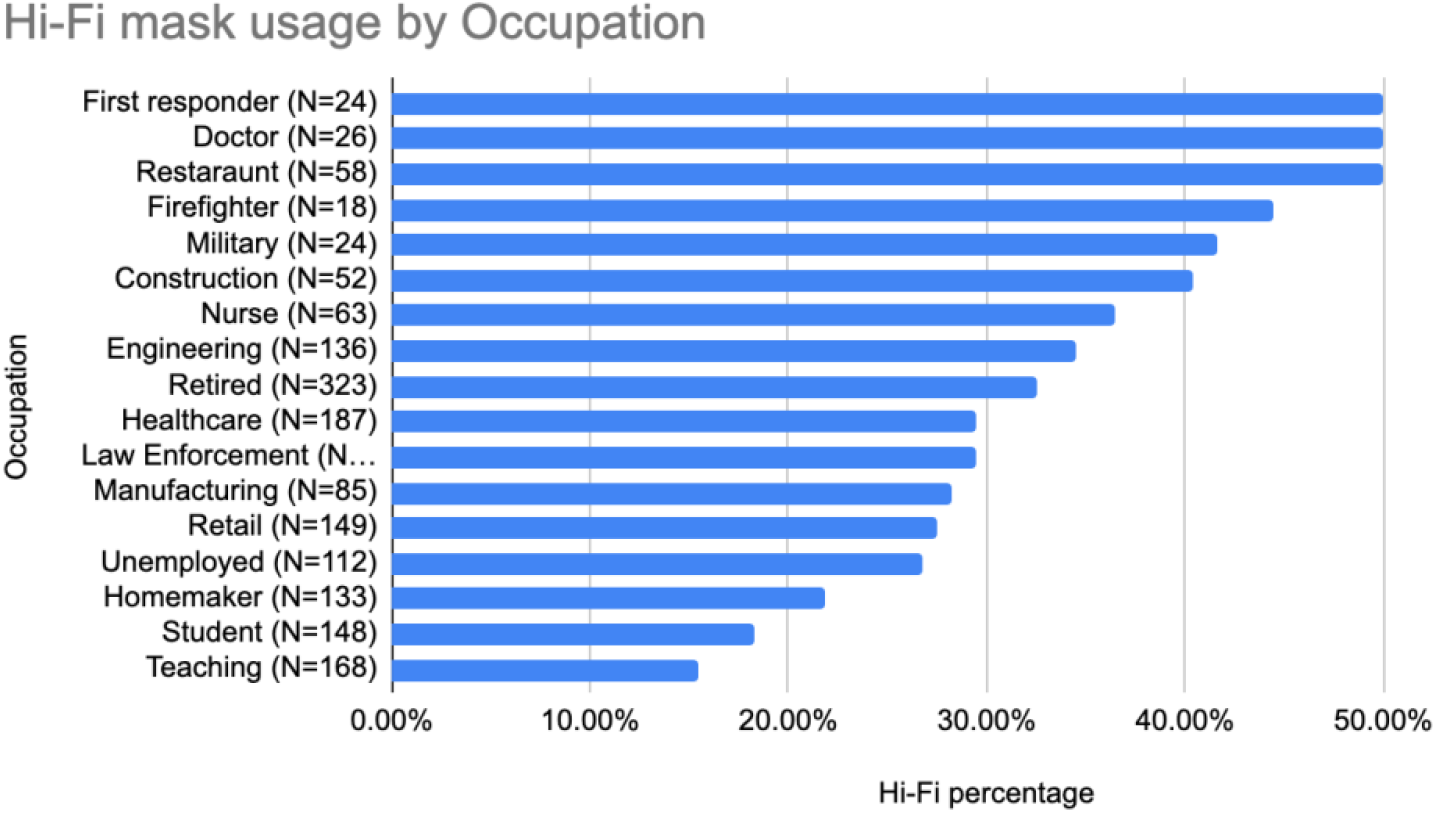
Percentage of US residents using Hi-Fi masks by occupation.

Respondents self-reported usage of their masks is shown in Figure 5. A majority of respondents reported using masks at least monthly. Nationally a third do so daily with a slightly higher percentage (42%) in the Pacific and Mountain regions.

**Figure 5:**
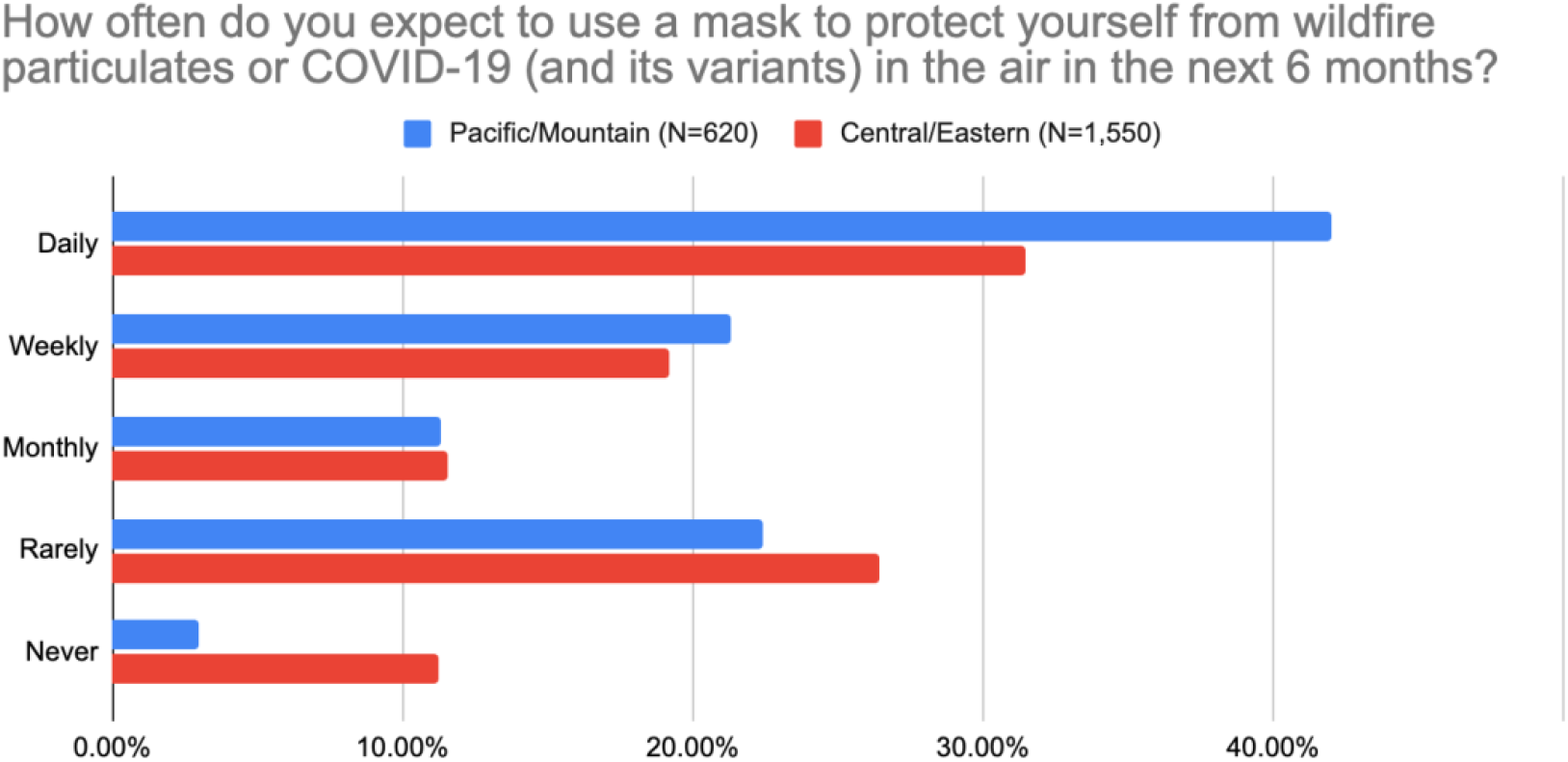
How often US residents use masks to protect from COVID-19 or wildfires.

### Less than half use air purifiers, and of them, less than half calculated the size needed

As shown in Figure 6, a greater percentage of respondents reported using air purifiers or HEPA air cleaners in the Pacific/Mountain zones (52%) than in the Central/Eastern zones (40%). However only 1 in 5 reported using these in their bedroom where they sleep (and likely spend significant amounts of time).

**Figure 6:**
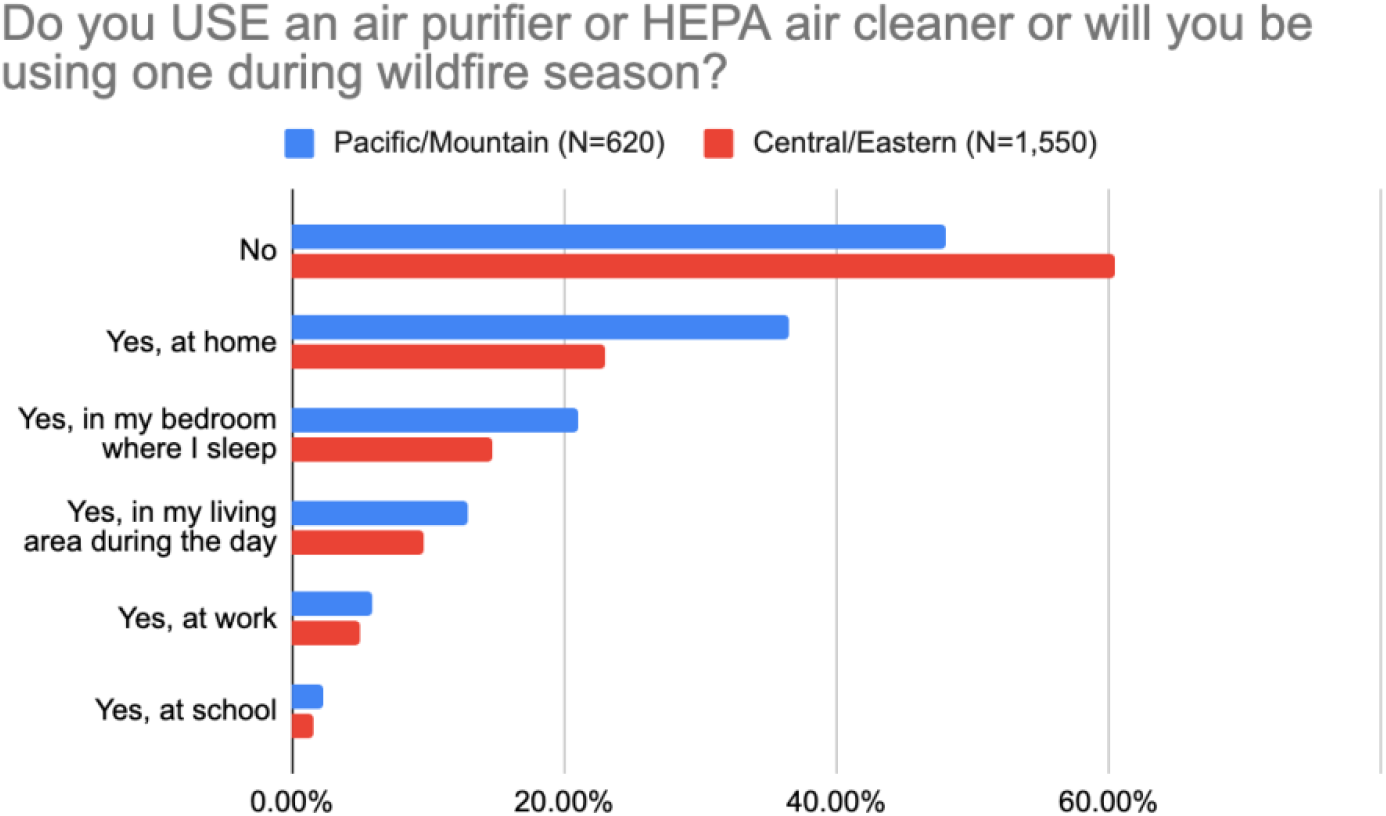
Where US residents use air purifiers or HEPA air cleaners.

As with Hi-Fi masks, higher than average use of air purifiers or HEPA air cleaners at home was reported by those in certain occupations (first responders, doctors, nurses, law enforcement, military, construction, firefighters, restaurant industry, and engineering) as shown in Figure 7.

**Figure 7:**
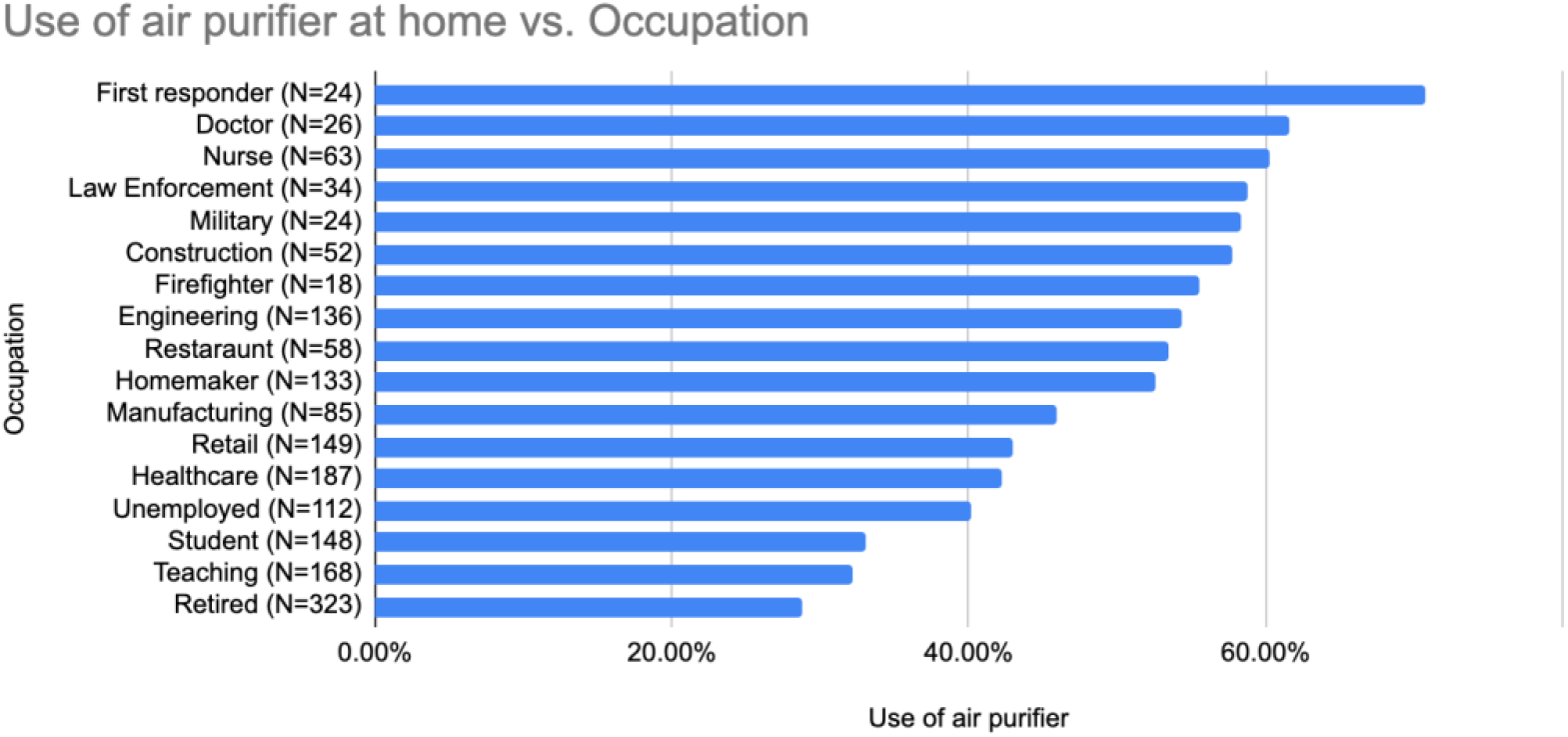
Use of air purifiers or HEPA air cleaners by occupation of US residents.

A common mistake is to choose a HEPA air cleaner that is too small for the size of the room it is used and ends up not having the intended effect. The correct size can be determined by a simple but perhaps tedious calculation involving the dimensions of the room and the clean air delivery rate (CADR). To choose the size of the air purifier or HEPA air cleaner used in their home the majority (51%) of respondents selected popular models or based their choice on advice from friends, colleagues, reviews, and articles. Only a minority did a calculation to determine the size needed in the room they use it in as shown in Figure 8 using either a calculator or by doing the calculation themselves.

**Figure 8:**
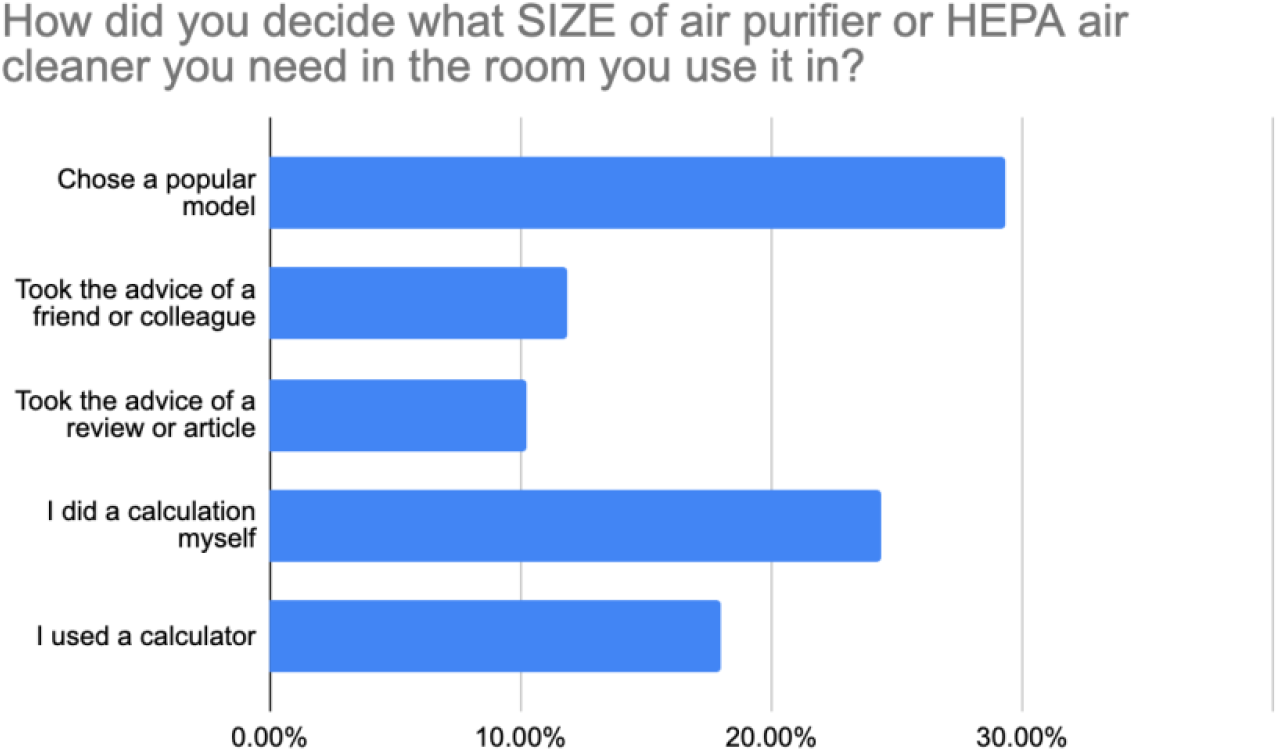
How US residents selected the size of their air purifiers or HEPA air cleaners.

**Figure 9:**
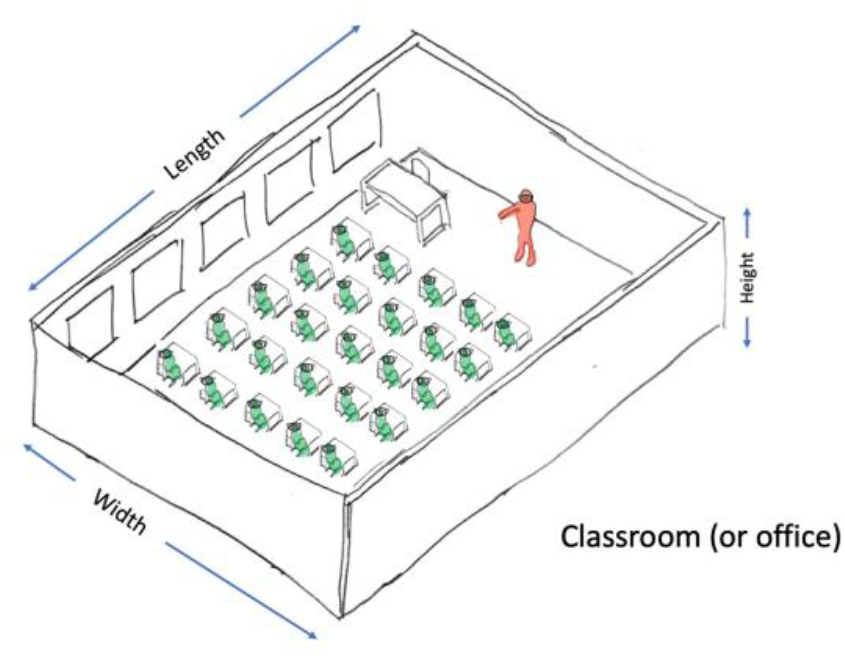
Room dimensions are needed to estimate the size of HEPA air cleaner needed.

## Discussion

In June and July of 2021, in our survey of 2,250 US residents, 66% are worried about inhaling COVID-19 and its variants whereas 52% are worried about toxic wildfire particles. In the mountain and pacific regions the latter rises to 73%. However, our survey also reveals that only a minority are using proven air filtering tools, or if they are using the right tools and some of them may also be doing so insufficiently. To protect from toxic wildfire particles or COVID variants that may arise, only 27% of US residents surveyed plan to use Hi-Fi masks and in the Pacific/Mountain zones only 21% plan to use air purifiers or HEPA air cleaners at home in their bedroom during wildfire season.

### HEPA air cleaners

Although fresh air ventilation from outside is useful to reduce aerosolized viral transmission, when the source of the toxic aerosol is outdoors during wildfire season access to fresh outdoor air is not always feasible. For such scenarios, the CDC recommends [25] portable HEPA cleaners including in classrooms [39]. Generally, one HEPA air cleaner is needed in each room.

Aerosols have been demonstrated to be reduced anywhere from 2x [26] to 4x [27] to 10x [28] compared to outdoors depending on how well the size of HEPA air cleaner is matched to the room. A common mistake is to use a HEPA air cleaner that is insufficient for the size of the room (office, classroom, etc.) and ends up being less effective if not ineffective.

Estimating the size of HEPA air cleaner needed is straightforward but can be tedious, and it depends on the volume of the room (length x width x height) and how many times per hour the air needs to be cleaned. Calculators to estimate the capacity (size) of the HEPA air cleaner needed in a room or set of rooms are available online [29] [30].

Particles containing Coronavirus accumulate in the air in shared spaces like classrooms or offices. HEPA air cleaners are useful to reduce this. The automatic fan sensor in most HEPA cleaners is designed to detect solid particles (e.g. from wildfires) but not typically the aerosols containing Coronavirus in the exhaled breath. It is therefore very important to override the “auto” setting and run the HEPA air cleaner’s fan on maximum speed (or if max speed is too noisy then the next one down) to get the air filtration benefit in such shared spaces. Also, it is important not to forget to remove the plastic cover on the filter inside a new HEPA air cleaner when it is first unpacked.

During the 2020 wildfire season in the Western United States there was an acute shortage of HEPA cleaners. 2021 may have more such shortages with a more severe fire season expected.

### Hi-Fi masks

A recent study [33] suggests Hi-Fi masks of sufficient quality are effective in preventing COVID-19 transmission when used appropriately. These same Hi-Fi masks also significantly reduce inhalation of toxic wildfire particles and the CDC recommends N95 masks [34] for this purpose. NIOSH (National Institute for Occupational Safety and Health) approval (e.g. N95, N99, P95, etc.) confirms a mask has been validated by the US government. In spite of supply constraints during the early days of the pandemic, Hi-Fi masks are now available without supply limitations such as N95 respirators and elastomeric N95 respirators (eN95) [31] [32] [35] [36] [37].

### Beyond COVID-19 and Wildfires

COVID-19 will not be the last nor worst-case respiratory virus with pandemic potential. Global pandemics from novel viruses may be accelerating. We saw Ebola in 2014, Zika in 2016, and Coronavirus in 2019. There are over 180 human viruses besides Covid-19, and on average two new species are added every year [16]. Some 94% of these are RNA viruses. RNA viruses are prone to rapid mutations because they lack a proofreading mechanism, and have rapid replication [17].

For example, could a lab turn measles-like dog virus (∼50% fatal) into a human virus? Consider a dog virus called Canine Distemper Virus (CDV) [18] which acts by first attacking the immune system (via SLAMF1=CD150) then attacks the nervous system (via PVRL4). In vitro, gain of function studies suggest this canine virus may be just 2-3 mutations away from infecting humans [19]. Mutations in viruses are routinely synthesized artificially as part of COVID-19 vaccine research in laboratories across the world [20].

Aerosolized threats from bacteria dispersal [21] motivated the development of a national sensor network called Biowatch [22]. The idea of a lightweight respirator to secure against biological attacks that requires low inhalation pressures, is comfortable to wear for prolonged periods, and will not interfere with vision, hearing, and communication was described in 1996 [23]. Today these are available off-the-shelf for low-cost in the form of Hi-Fi masks (e.g. elastomeric N95).

The health risks from the COVID-19 delta variant and wildfire pollution in the short-term, and the long-term threats of airborne pandemics and biological attacks can be reduced by use of aerosol precautions.

## Data Availability

Due to the nature of this survey data collection method, participants of this survey did not agree for individual response data to be shared publicly, so supporting data is not available.

